# Acute effects of static stretching to hamstrings on spatiotemporal variables and leg kinematics in late swing at maximal sprint speed phase and its relationship with Nordic hamstring strength

**DOI:** 10.1101/2023.03.06.23286888

**Authors:** Yusuke Ozaki, Takeshi Ueda

## Abstract

This study aimed to determine the acute effects of static stretching of the hamstrings on maximal sprint speed and its spatiotemporal variables, and lower limb kinematics during the late swing phase, and its relationship with Nordic hamstring strength. Sixteen healthy male college sprinters were asked to sprint 80 m without static stretching and with static stretching of the hamstrings for 120 s per leg before the sprint, and both conditions were counterbalanced. The knee flexion peak force and torque were measured using the Nordic hamstring. The differences between no static stretch and static stretch, and their relationship with Nordic hamstring strength were investigated. The results showed that the touchdown distance and support time increased, and flight distance decreased in under static stretch conditions with a decrease in maximal sprint speed. Moreover, under static stretch conditions, the angular velocity of knee extension at contralateral release was lower, while the theoretical hamstring length (difference between knee angle and hip angle) at ipsilateral touchdown was greater. In addition, the lower the peak force and torque of the Nordic hamstring, the more significant the decrease in maximal sprint speed, increase in support time, decrease in flight distance, and decrease in peak angular velocity of hip extension at static stretch. Furthermore, the more significant the decrease in maximal sprint speed at static stretch, the smaller the peak theoretical hamstring length at the no static stretch. Therefore, it is suggested that long-term static stretching immediately before sprinting in sprinters with poor Nordic hamstring strength and low hamstring compliance during the late swing phase may induce unfavorable kinematics to prevent hamstring strain injury and maximal sprint speed reduction.

## Introduction

Maximal sprint speed (MSS) is an important motor skill related to performance in sports that require high-speed sprinting, such as track and field, soccer, and rugby [1, 2]. During high-speed sprints, the incidence of hamstring strain injuries (HSI) is increased [3, 4]. HSI has a significant impact on an athlete’s career because of the high re-injury rate and prolonged withdrawal from competition [5, 6]. Therefore, presenting a scientific perspective on HSI prevention is an important research topic for improving the efficiency of training for high-speed running ability improvement and high performance in competitions.

During the late swing phase (the phase from maximal hip flexion to ground contact) in this MSS phase, the hamstrings are more active because of the eccentric deceleration of hip flexion and knee extension movements [7]. In addition, the length and strain of the hamstring muscle-tendon units that contribute to hip extension and knee flexion reach their peak [8, 9]. Therefore, HSI is thought to occur more frequently during the late swing phase [10–15].

HSI risk is associated with a short hamstring fascicle length [16, 17] and poor eccentric muscle strength [16, 18–21], and training protocols have been developed to reduce HSI risk [22]. In particular, hamstring eccentric training has become popular as a typical training to reduce the risk of HSI because it can lengthen the fascicle length, improve eccentric strength with a longer hamstring, and strengthen the ability of the hamstring to resist excessive muscle stretching [23–30].

Hamstring muscle-tendon unit stiffness has also been reported as an HSI risk [5, 31, 32]. This is because the high compliance of the muscle-tendon units contributes to energy absorption during the muscle-tendon unit extension [33]. The stiffness of the muscle-tendon units is partially due to muscle stiffness [34, 35]. Therefore, static stretching before sport-specific training that acutely improves the compliance of muscle-tendon units is recommended and is frequently performed among track and field sprinters, where HSI often occurs [6, 36, 37]. Recent studies have shown that static stretching of the hamstrings improves eccentric muscle strength at longer muscle lengths, highlighting the benefits of static stretching in HSI prevention [29, 30, 38, 39]. Thus, while the injury-preventive effects of static stretching were previously questioned, the value of using static stretching has recently been reconsidered for certain injuries [40].

However, static stretching impairs the subsequent explosive strength [41, 42]. In sprinting, static stretching immediately prior to exercise, including the hamstring, impairs performance from 5–40 m [42]. In contrast, Kistler et al. [43], reported that in the 100 m sprint, static stretching acutely impaired sprint speed at 20–40 m but did not further impair sprint speed at 60 m thereafter. The MSS phase in the 100 m sprint is approximately 40–60 m, which requires a higher eccentric torque in hip extension and knee flexion and higher activation levels in the hamstrings than in the acceleration phase [44–46]. Nevertheless, if MSS is not impaired by static stretching of the hamstrings, then static stretching to increase hamstring compliance is a beneficial means of HSI prevention in athletes’ competitions and training where MSS is required.

However, static stretching of the hamstrings decreases the hamstring‒quadriceps ratio [47], which is an HSI risk [21, 48, 49]. Therefore, relative hamstring strength reduction may promote hamstring muscle-tendon unit lengthening (that is, excessive hip flexion and knee extension) during the late swing phase, resulting in running kinematics that is unfavorable for HSI prevention [50, 51]. In addition, hamstring muscle volume is an important indicator of sprint performance [52]. Based on the above, we cannot dismiss the possibility that static stretching of the hamstring alone may produce undesirable kinematics and changes in spatiotemporal variables for HSI risk reduction and MSS improvement. In contrast, because many studies have reported minimal negative effects of static stretching in highly trained elite athletes [53–58], this level of performance and muscle strength may influence the acute effects of static stretching.

However, no studies have investigated the acute effects of static stretching of the hamstring alone on the MSS phase, and the relationship between spatiotemporal variables and kinematics during sprinting, performance level, and muscle strength level. These findings may be useful as guidelines for athletes, coaches, and physical therapists concerned with achieving MSS.

Therefore, this study aimed to determine the acute effects of static hamstring stretching on MSS and its spatiotemporal variables and kinematics during the late swing phase, and its relationship with hamstring strength and performance levels. The hypotheses of this study are as follows: (1) Static stretching of the hamstrings prior to sprinting acutely decreases sprint speed by decreasing step rate and increasing knee joint angle during the late swing phase. (2) The lower the hamstring strength, the greater the decrease in sprint speed owing to static stretching.

## Materials and methods

### Participants and experimental procedures

Participants were 16 collegiate sprinters (100 m personal record [PR]; 11.25±0.50 sec; Height: 173.22±5.97 cm; Mass: 68.00±5.81 kg, age: 21.56±2.25 years). In addition, two participants with HSI injuries from the previous year were included in this study. The two HIS injury dates were 4 months earlier, and they had already returned to the competition. All participants were recruited during the period September-October 2022. All participants were informed of the experimental procedures and risks, and they provided written informed consent prior to participation in the study. The experiment was conducted without discomfort to the participants and in accordance with the principles of the Declaration of Helsinki. The local ethics review board (Graduate School of Humanities and Social Sciences, Hiroshima University) approved the experimental protocol (approval number: 2021063). Measurements were performed in a within-subject experimental design and were counterbalanced in the no static stretch (NS) and static stretch (SS) conditions. NS and SS measurements were taken at least 48 h apart. Participants performed a designated warm-up protocol that included 800-m running, dynamic stretching with hurdles, sprint drills with skipping, three 50-m sprints with a gradual increase in speed with each sprint, and a 30-m start dash with a starting block. Two 80-meter sprints with starting blocks were performed. Considering the effects of fatigue and the residual effect of static stretching (in SS trials) is 10–20 min [59], a 10-min rest period was provided between trials. The SS group performed a static stretching protocol prior to the first 80-m sprint trial and then completed the first 80-m sprint trial within 5 min. After static stretching, the athletes were allowed to practice one or two block clearances before starting the 80-m sprint trial. SS was performed by passive static straight leg raising to stretch the hamstring. The participant was placed supine on the mat, the contralateral hip and knee were stabilized in full extension, and the leg to be stretched was pushed by the investigator until the participant complained of discomfort. The knee was maintained at full extension and maximum hip flexion. A brief stretch of approximately 20 s is insufficient to decrease the passive stiffness of the muscle-tendon unit [60–62]. Therefore, the process of stretching the legs was continued for 30 s and repeated four times for each leg with a 10-s rest between stretches [39]. The same spike shoes were worn for both the NS and SS in the 80 m sprint trials, and the starting block placement was similar between the two conditions.

The Nordic hamstring (NDH) was used to measure hamstring strength. The NDH was adopted because of its proven HSI risk reduction effectiveness, simplicity, and practicality [63]. NDH measurements were performed within 2 weeks of the 80 m sprint trial for the two conditions.

Prior to NDH measurements, two sets of three NDH were performed as a familiarization session. The NDH measurements were then taken after a minimum of 72 h. Prior to the measurements, the participants performed a specified warm-up that included 5 min of pedaling (2 kp, 60 rpm) on a stationary aerobike (PowerMax V II, COMBI, Japan), dynamic stretching with hurdles, and three sets of five squat jumps. To measure NDH, a dedicated NDH strength instrument (N3, Easytech, Italy) with a pressure gauge attached to the heel was used. The participants’ ankles were fully immobilized, holding full hip extension, and the elbows bent. The participant then leaned forward at the slowest possible motion with the elbows on the body side and hands open [64]. Once the participants lost control of their forward lean, they were allowed to increase their speed and fall forward. NDH was measured three times with a 2-min rest in between.

### Experimental setup

A high-speed camera (EX-100F, CASIO, Tokyo, Japan) was used to capture video at 240 frames/s for the 52.5–60 m section and the MSS section of the 80 m sprint test [65]. The camera was positioned 20 m to the right of the center of the analysis section, with the optical axis perpendicular to the direction of the sprint. Prior to trial filming, markers were placed in a rectangular pattern on the ground in the center of the filming area at intervals of 1.2 m in the lateral direction and 2.5 m in the direction of the sprint. To calibrate the aspect ratio, images were taken with a 2-m calibration pole held vertically and horizontally at the center of the marker.

### Data analysis

Video images were captured on a personal computer, and a skilled examiner manually digitized 21 body feature points at 120 Hz using a video motion analysis system (Frame-DIAS V, DKH Retail Limited, Cheltenham, United Kingdom). The participants wore elastic black tights over their entire body, and 30 white markers were attached to each participant’s body and shoes to ensure that body feature points could be identified. The markers were positioned at the top of the head, upper margin of the sternum, right and left tragion, acromion, greater trochanters, medial and lateral epicondyles, styloid process of the radius and ulna, head of the third metacarpal bone, medial and lateral epicondyle of the femur, medial and lateral malleolus, and heel and toe (on shoes). All markers were applied by the same examiner to ensure consistency in the marker location. The participants’ measurement variables were calculated using a two-dimensional four-point real-length conversion method based on the calibrations. Center of mass (CM) coordinates were obtained using the inertial parameters of the 14-segment model reported by Ae [66]. All trials were digitized from 10 frames before the left foot touchdown until 10 frames after the next left foot touchdown.

Based on the digitized data, actual coordinate values were obtained for each trial, with the x-axis for the sprint direction and the y-axis for the vertical direction. The obtained two-dimensional coordinates were smoothed for each coordinate component of each analysis point using a Butterworth digital filter, after determining the optimal cutoff frequency using the residual analysis method [67]. The actual cutoff frequencies in the sprint and vertical directions were in the ranges of 8–14 Hz and 4–15 Hz, respectively.

### Variables

Fig 1 shows the definitions of the variables calculated in this study. The trial with the highest MSS among the two trials was used for the analysis.

**Fig 1.**
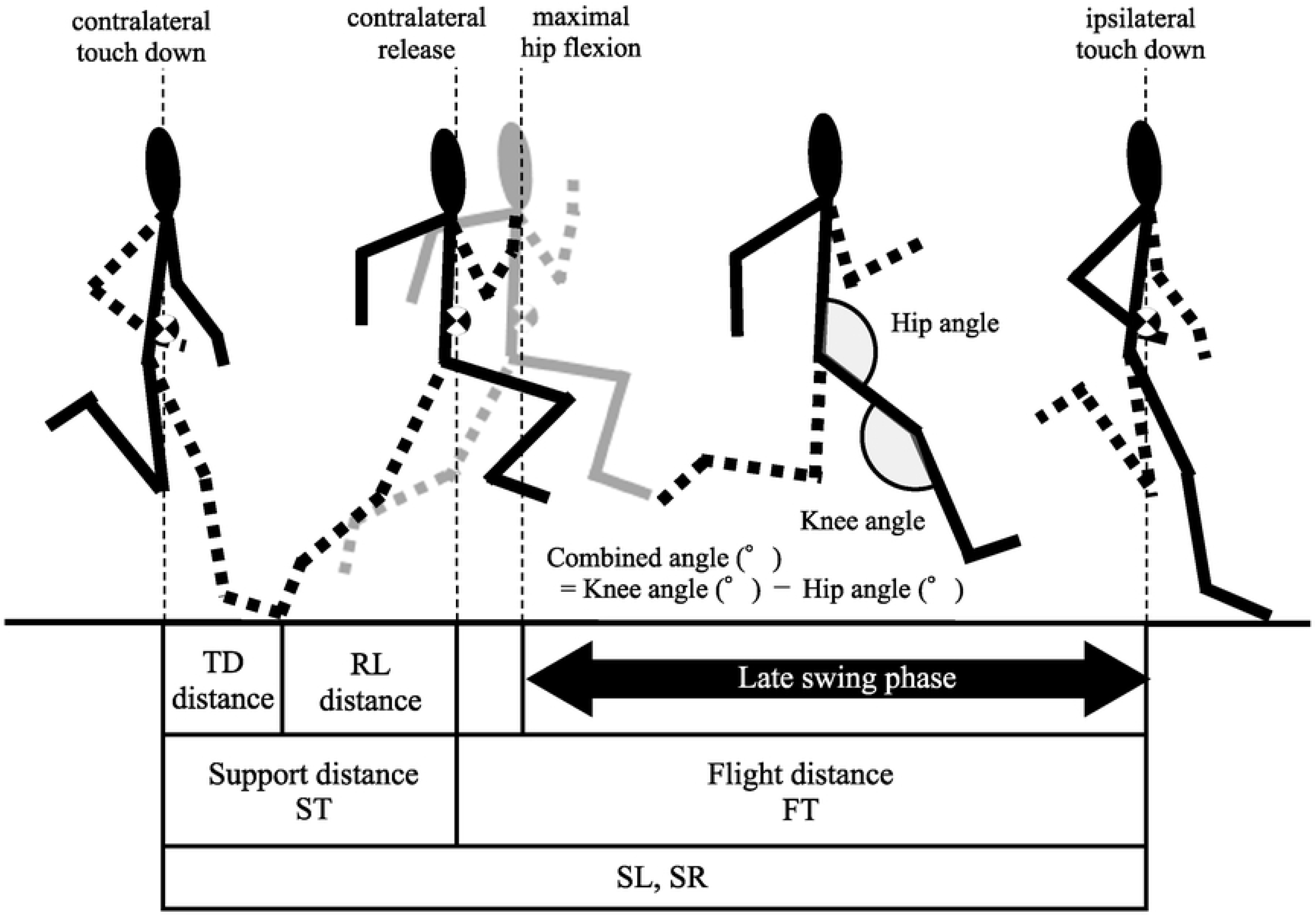
Definition of calculated variables in this study. TD distance, touchdown distance; RL distance, release distance; ST, support time; FT, flight time; SL, step length; SR, step rate.

#### Spatiotemporal variables

- **Maximal sprinting speed (MSS [m·s^-1^])**: The average horizontal speed from the first left foot touch-down to the next left foot touchdown.
- **Step length (SL [m])**: Horizontal displacement of the center of mass divided by 2 in two steps from the first left foot touching down to the next left foot touching down.
- **Step frequency (SR [step·s^-1^]])**: Reciprocal of the time required for two steps from the first left foot touchdown to the next left foot touchdown divided by 2.
- **Touch-down distance (TD-distance [m])**: Horizontal distance between the toe and center of mass at the moment the toe touches down.
- **Release distance (RL-distance [m])**: Horizontal distance between the toe and center of mass at the moment of the toe release.
- **Support time (ST [s])**: Time required from the moment the toe touches down and the moment the toe is released.
- **Flight time (FT [s])**: the time required from the moment of toe release to the moment of opposite-toe touchdown.
- **Support distance (m)**: Horizontal displacement of the center of mass from the moment the toe touches down to the moment it is released.
- **Flight distance (m)**: Horizontal displacement of the center of mass from the moment of the toe release to the moment of touchdown of the opposite toe.

The average value in the two steps during the first left-toe touchdown and the subsequent left-toe touchdown was used to maintain the reliability of the values in TD distance, RL distance, ST, FT, Support distance, and flight distance.

#### Kinematic variables

Kinematic analysis in this study was performed from the moment of the release of the left toe (contralateral leg) to the moment of the touchdown of the right toe (ipsilateral leg).

This is because it corresponds to the late swing phase, which has the highest hamstring muscle-tendon unit elongation, strain, and HIS [10–15].

- **Hip angle (°), Hip angular velocity (°·s^-1^)**: angle and angular velocity formed by the line connecting the upper margin of the sternum, right greater trochanter, and right knee.
- **Knee angle (°), Knee angular velocity (°·s^-1^)**: Angular velocity formed by the line connecting the right greater trochanter, right knee, and right ankle.
- **Combined angle (°)**, angle obtained by subtracting the hip angle from the knee joint angle. The combined angle is a measure of the theoretical hamstring muscle-tendon unit length [50].

For statisticcoe release, peak values during the late swing phase, and ipsilateral toe touchdown were used.

To assess measurement repeatability, the same investigator analyzed these variables twice for 10 participants, spaced at least 48 h apart. The intraclass correlation coefficient for the two analyses averaged 0.893 ± 0.055 (lowest 0.774, highest 0.972). The smallest coefficient of variation was the MSS (0.4 %), and the largest was the combined angle at RL (9.4%). These were recognized as having good intrarater reliability. A series of mixed-model analysis of variances were also performed to examine the effects of the measurement date and order of NS and SS. The results showed no significant measurement date effects, order effects, or interactions for any of the variables.

### NDH strength

- **NDH peak force (N·kg^-1^)**: The average of the measured peak force of the left and right feet divided by the body weight.
- **NDH peak torque (Nm·kg^-1^)** : Peak force multiplied by the lower leg length of the participant.

The NDH peak force and torque intraclass correlation coefficients for the three measurements were 0.904 and 0.912, respectively. The coefficient of variations were 4.02 ± 2.06 % in both cases. Thus, they were accepted as having good inter-measurement reliability.

The trial with the highest NDH peak force for three measurements was used in the analysis.

### Statistical analyses

Paired t-tests were used to test for differences in NS and SS variables. Effect sizes were categorized as small (0.20–0.49), medium (0.50–0.79), large (0.80–1.19), very large (1.2–1.99), or huge (>2.0) according to Cohen’s effect size [68]. Pearson’s product-rate correlation coefficient was calculated to examine the relationship between NDH strength and the amount of change in variables from NS to SS. Statistical analyses were performed using statistical processing software (IBM SPSS Statistics v20.0, IBM, Japan). Each variable was presented as mean ± standard deviation (SD). The amount of change (Δ) in variables from NS to SS is shown as mean ± standard error (SE). The statistical significance level was set at p < 5%.

## Results

Table 1 shows the values of the spatiotemporal variables in the NS and SS conditions, and the differences between the conditions. The MSS was lower in 11 of 16 participants in the SS condition and higher in five participants in the SS condition (mean ±SD; -2.47±4.37 %).

**Table 1.**
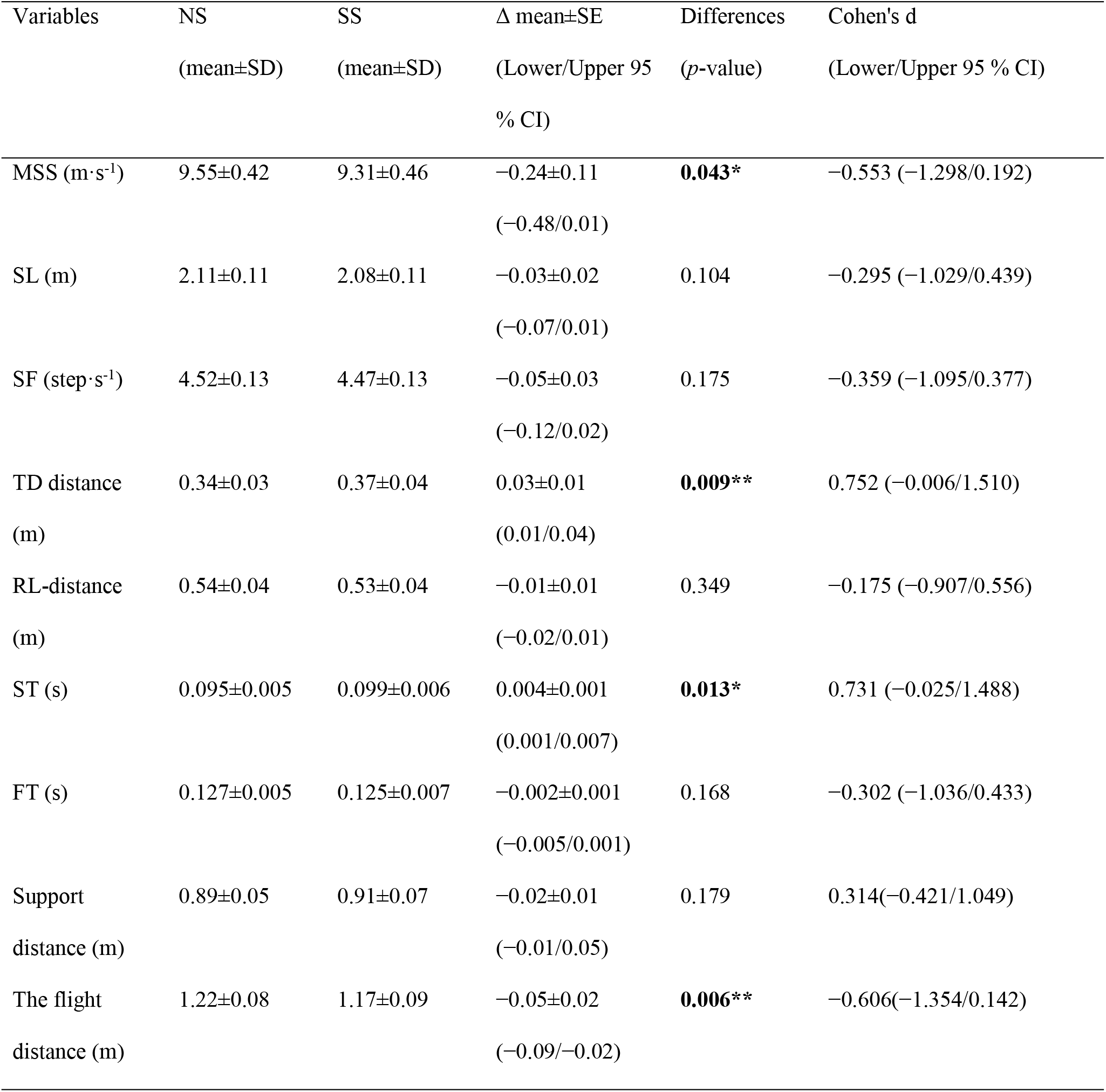
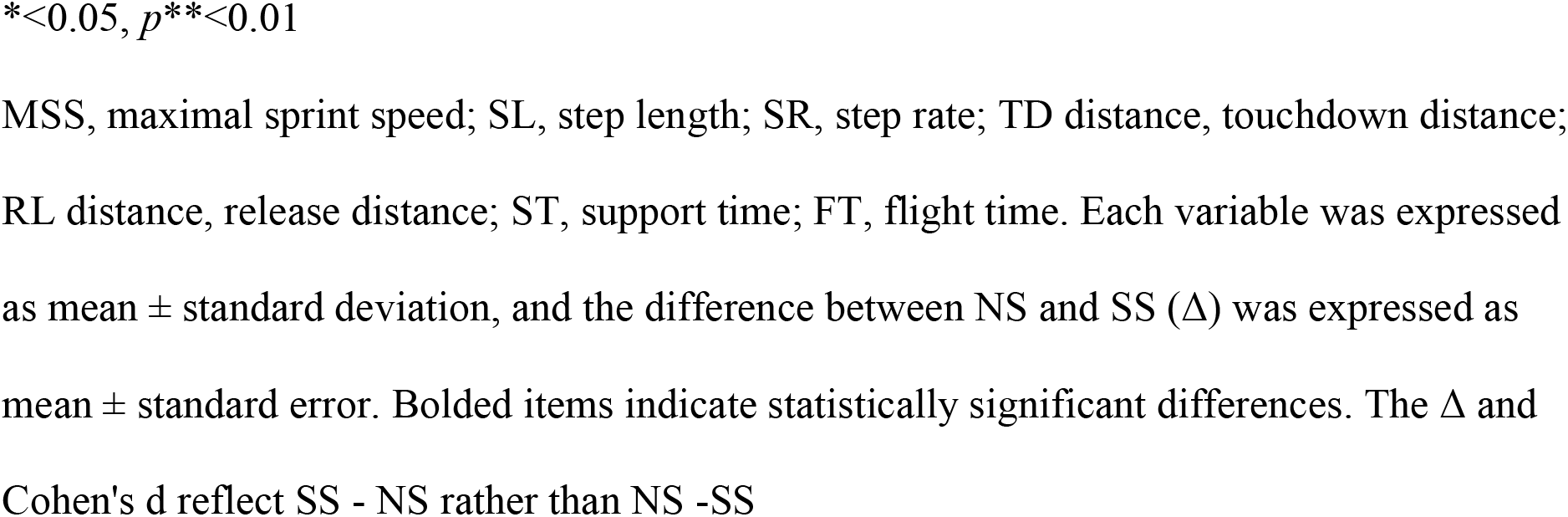
Values and differences of spatiotemporal variables in NS and SS conditions.

| Variables | NS<br>(mean±SD) | SS<br>(mean±SD) | $\Delta$ mean±SE<br>(Lower/Upper 95<br>% CI) | Differences<br>(*p*-value) | Cohen's d<br>(Lower/Upper 95 % CI) |
| --- | --- | --- | --- | --- | --- |
| MSS (m·s<sup>-1</sup>) | 9.55±0.42 | 9.31±0.46 | -0.24±0.11<br>(-0.48/0.01) | **0.043\*** | -0.553 (-1.298/0.192) |
| SL (m) | 2.11±0.11 | 2.08±0.11 | -0.03±0.02<br>(-0.07/0.01) | 0.104 | -0.295 (-1.029/0.439) |
| SF (step·s<sup>-1</sup>) | 4.52±0.13 | 4.47±0.13 | -0.05±0.03<br>(-0.12/0.02) | 0.175 | -0.359 (-1.095/0.377) |
| TD distance<br>(m) | 0.34±0.03 | 0.37±0.04 | 0.03±0.01<br>(0.01/0.04) | **0.009\*\*** | 0.752 (-0.006/1.510) |
| RL-distance<br>(m) | 0.54±0.04 | 0.53±0.04 | -0.01±0.01<br>(-0.02/0.01) | 0.349 | -0.175 (-0.907/0.556) |
| ST (s) | 0.095±0.005 | 0.099±0.006 | 0.004±0.001<br>(0.001/0.007) | **0.013\*** | 0.731 (-0.025/1.488) |
| FT (s) | 0.127±0.005 | 0.125±0.007 | -0.002±0.001<br>(-0.005/0.001) | 0.168 | -0.302 (-1.036/0.433) |
| Support<br>distance (m) | 0.89±0.05 | 0.91±0.07 | -0.02±0.01<br>(-0.01/0.05) | 0.179 | 0.314(-0.421/1.049) |
| The flight<br>distance (m) | 1.22±0.08 | 1.17±0.09 | -0.05±0.02<br>(-0.09/-0.02) | **0.006\*\*** | -0.606(-1.354/0.142) |

\* $<0.05$ , $p^{**}<0.01$
MSS, maximal sprint speed; SL, step length; SR, step rate; TD distance, touchdown distance; RL distance, release distance; ST, support time; FT, flight time. Each variable was expressed as mean ± standard deviation, and the difference between NS and SS ( $\Delta$ ) was expressed as mean ± standard error. Bolded items indicate statistically significant differences. The $\Delta$ and Cohen's d reflect SS - NS rather than NS -SS

Some of them exhibited a velocity decrease of 1.37 m/s. SS tended to have a smaller hip angle, larger knee angle, and larger combined angle than NS during the late swing phase toward ipsilateral touchdown (Fig 2).

**Fig 2.**
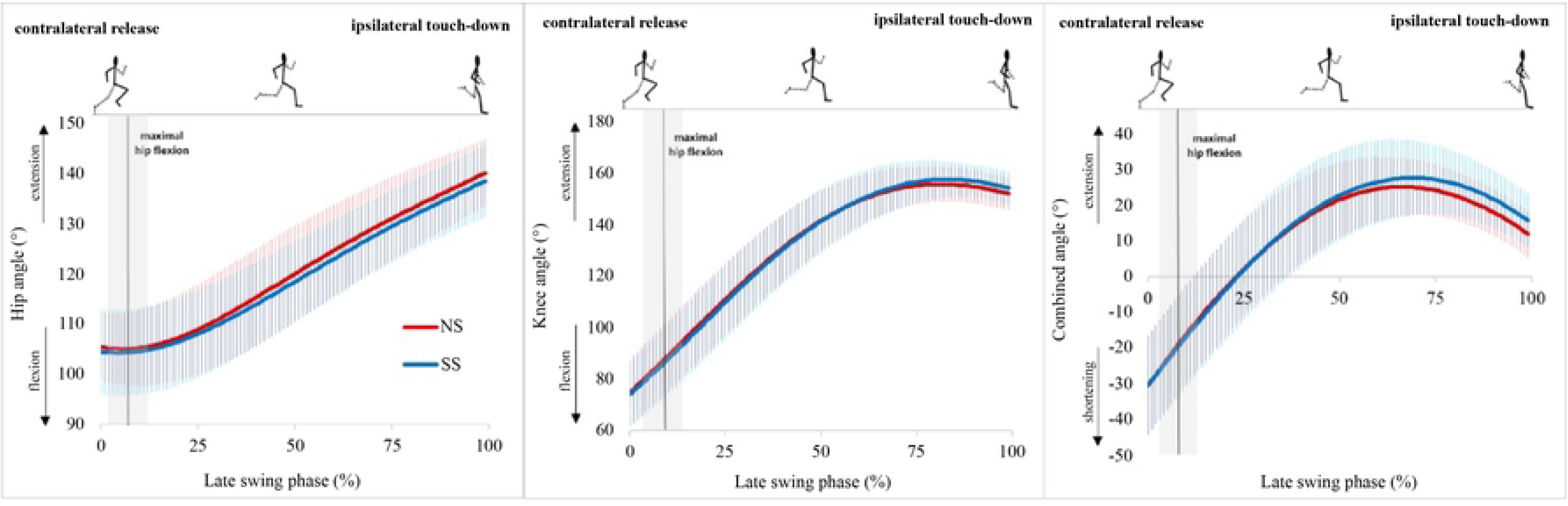
Changes in the hip angle, knee angle, and combined angle during the late swing phase.

In particular, the combined angle was significantly larger in SS than in NS at the ipsilateral touchdown moment (Table 2).

**Table 2.** Values and differences of kinematic variables in NS and SS conditions.

| Variables | NS<br>(mean±SD) | SS<br>(mean±SD) | $\Delta$ mean±SE<br>(Lower/Upper 95<br>% CI) | Differences<br>($p$-value) | Cohen's d<br>(Lower/Upper 95<br>% CI) |
| --- | --- | --- | --- | --- | --- |
| Hip angle at RL ($^{\circ}$) | 105.4±7.01 | 104.41±8.72 | -1.00±1.38<br>(-3.93/1.94) | 0.481 | -0.127(-0.857/0.604) |
| Peak hip flexion<br>angle ($^{\circ}$) | 104.53±7.17 | 103.8±8.65 | -0.73±1.41<br>(-3.73/2.28) | 0.614 | -0.092(-0.822/0.638) |
| Hip angle at TD (°) | 140.15±6.83 | 138.58±7.51 | -1.57±1.24<br>(-4.21/1.08) | 0.226 | -0.219(-0.952/0.5<br>13) |
| Knee angle at RL (°) | 74.84±13.40 | 74.07±13.21 | -0.77±1.43<br>(-3.81/2.27) | 0.597 | -0.058(-0.788/0.6<br>72) |
| Peak knee extension<br>angle (°) | 157.1±6.79 | 159.12±6.89 | 2.02±1.07<br>(-0.26/4.30) | 0.078 | 0.298(-0.437/1.03<br>2) |
| Knee angle at TD (°) | 152.11±6.55 | 154.27±6.87 | 2.16±1.29<br>(-0.60/4.91) | 0.116 | 0.323(-0.412/1.05<br>8) |
| Combined angle at<br>RL (°) | -30.56±14.37 | -30.34±14.52 | 0.23±1.85<br>(-3.71/4.16) | 0.905 | 0.016(-0.714/0.74<br>5) |
| Peak combined angle<br>(°) | 26.28±8.31 | 28.42±10.05 | 2.14±1.22<br>(-0.46/4.74) | 0.100 | 0.234(-0.499/0.96<br>6) |
| Combined angle at<br>TD (°) | 11.97±6.73 | 15.69±7.80 | 3.72±1.46(0.60/6<br>.85) | **0.023\*** | 0.514(-0.229/1.25<br>7) |
| Hip angular velocity<br>at RL (°·s<sup>-1</sup>) | -98.8±151.48 | -55.73±155.97 | 43.07±20.72<br>(-1.10/87.24) | 0.055 | 0.282(-0.452/1.01<br>6) |
| Peak hip extension<br>velocity (°·s<sup>-1</sup>) | 434.54±90.11 | 423.17±67 | -11.38±21.61<br>(-57.44/34.68) | 0.606 | -0.144(-0.875/0.5<br>86) |
| Hip angular velocity<br>at TD (°·s<sup>-1</sup>) | 302.25±113.73 | 300.14±125.63 | -2.11±42.20<br>(-92.06/87.83) | 0.961 | -0.018(-0.747/0.7<br>12) |
| Knee angular<br>velocity at RL (°·s<sup>-1</sup>) | 1086.72±129.71 | 1042.06±174.46 | -44.66±20.57<br>(-88.50/-0.83) | **0.046\*** | -0.293(-1.027/0.4<br>42) |
| Peak knee extension<br>velocity (°·s<sup>-1</sup>) | 1200.53±119.22 | 1224.94±153.55 | 24.41±25.89<br>(-30.77/79.59) | 0.361 | 0.179(-0.053/0.91<br>0) |
| Knee angular<br>velocity at TD (°·s<sup>-1</sup>) | -287.88±130.59 | -308.04±201.46 | -20.16±41.45<br>(-108.51/68.20) | 0.634 | -0.120(-0.850/0.6<br>11) |

*p*\*<0.05
RL, Contralateral release; TD, ipsilateral touchdown. Items in bold are those with significant correlations. Δ and Cohen's d reflect SS-NS rather than NS.

Hip angular velocity was smaller in NS for contralateral release and tended to be higher in NS than in SS by approximately 25 % to 50 % (Fig 3, Table 2).

**Fig 3.**
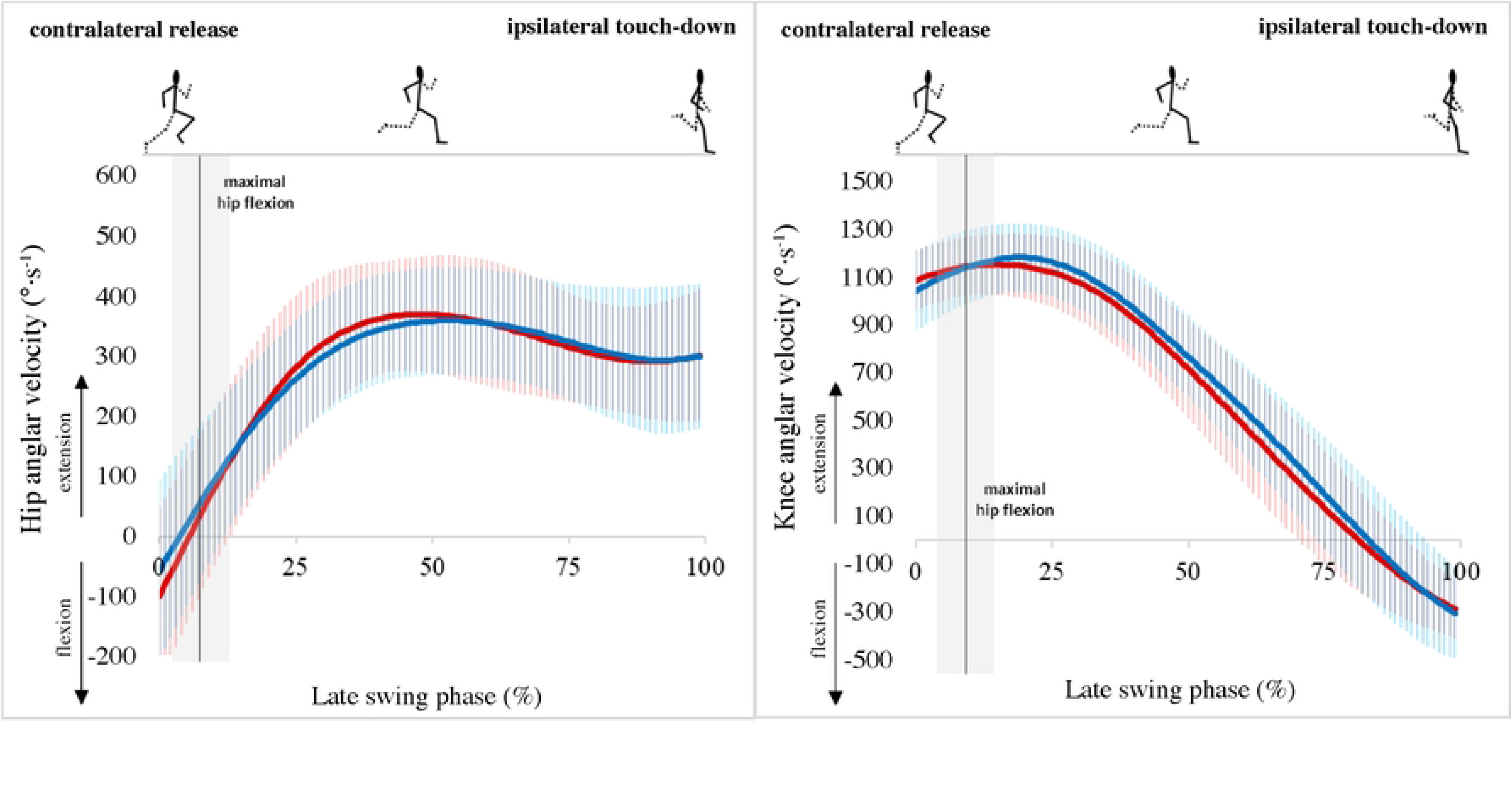
Changes in hip and knee angular velocities during the late swing phase.

During the contralateral release, NS had significantly higher knee angular velocity than SS, and although not significantly, SS had higher knee angular velocity than NS up to around the 90th percentile (Fig 3, Table 2).

The measured NDH peak force was 266.06 ± 50.41 N (body mass ratio; 3.95 ± 0.87 N kg^-1^) and NDH peak torque was 116.70 ± 22.09 Nm (body mass ratio; 1.73 ± 0.37 Nm kg^-1^). Table 3 presents the relationship between the amount of change in each variable across the conditions and NDH strength. There was a significant correlation between ΔMSS and NDH peak force (r=0.532, p<0.05) and peak torque (r=0.510, p<0.05).

**Table 3.** Relationship between the amount of change in each variable in NS and SS and NDH strength.

| Variables | NDH-peak-force (N/kg) |  | NDH-peak-torque (Nm/kg) |  |
| --- | --- | --- | --- | --- |
|  | Correlation coefficient<br>(Lower/Upper 95 % CI) | *p*-value | Correlation coefficient<br>(Lower/Upper 95 % CI) | *p*-value |
| $\Delta\text{MSS}$ ($\text{m}\cdot\text{s}^{-1}$) | 0.532 (0.049/0.813) | **0.034\*** | 0.510<br>(0.019/0.803) | **0.044\*** |
| $\Delta\text{SL}$ (m) | 0.380 (-0.143/0.737) | 0.147 | 0.372<br>(-0.152/0.732) | 0.156 |
| ΔSF (step·s<sup>-1</sup>) | 0.391(−0.130/0.743) | 0.135 | 0.367<br>(−0.157/0.730) | 0.162 |
| ΔTD distance (m) | −0.26 (−0.670/0.270) | 0.330 | −0.209<br>(−0.639/0.319) | 0.436 |
| ΔRL-distance (m) | −0.093 (−0.563/0.422) | 0.732 | −0.088<br>(−0.559/0.426) | 0.746 |
| ΔST (s) | −0.605 (−0.847/−0.156) | **0.013\*** | −0.549<br>(−0.821/−0.073) | **0.028\*** |
| ΔFT (s) | 0.203 (−0.325/0.635) | 0.450 | 0.171<br>(−0.354/0.615) | 0.526 |
| ΔSupport distance (m) | −0.222 (−0.646/0.308) | 0.409 | −0.171<br>(−0.614/0.355) | 0.527 |
| ΔFlight distance (m) | 0.623 (0.184/0.855) | **0.010\*\*** | 0.571<br>(0.106/0.832) | **0.021\*** |
| ΔHip angle at RL (°) | −0.291 (−0.688/0.239) | 0.274 | −0.285<br>(−0.684/0.246) | 0.286 |
| ΔPeak hip flexion<br>angle (°) | −0.323 (−0.706/0.205) | 0.222 | −0.321<br>(−0.704/0.208) | 0.226 |
| ΔHip angle at TD (°) | 0.163 (−0.362/0.610) | 0.546 | 0.199<br>(−0.329/0.632) | 0.459 |
| ΔKnee angle at RL (°) | 0.207 (−0.322/0.637) | 0.442 | 0.119<br>(−0.400/0.581) | 0.660 |
| ΔPeak knee extension<br>angle (°) | 0.186 (−0.342/0.624) | 0.491 | 0.201<br>(−0.327/0.634) | 0.455 |
| ΔKnee angle at TD ($^{\circ}$) | -0.076 (-0.551/0.436) | 0.780 | -0.050<br>(-0.532/0.457) | 0.854 |
| ΔCombined angle at<br>RL ($^{\circ}$) | 0.377 (-0.146/0.736) | 0.150 | 0.305<br>(-0.225/0.695) | 0.251 |
| ΔPeak combined<br>angle ($^{\circ}$) | -0.120 (-0.581/0.399) | 0.657 | -0.153<br>(-0.603/0.371) | 0.571 |
| ΔCombined angle at<br>TD ($^{\circ}$) | -0.205 (-0.636/0.324) | 0.447 | -0.212<br>(-0.641/0.317) | 0.430 |
| ΔHip angular velocity<br>at RL ($^{\circ}\cdot s^{-1}$) | 0.146 (-0.378/0.598) | 0.591 | 0.054<br>(-0.454/0.536) | 0.841 |
| ΔPeak hip extension<br>velocity ($^{\circ}\cdot s^{-1}$) | 0.565 (0.096/0.829) | **0.023\*** | 0.626<br>(0.190/0.856) | **0.009\*\*** |
| ΔHip angular velocity<br>at TD ($^{\circ}\cdot s^{-1}$) | -0.099 (-0.567/0.417) | 0.715 | -0.106<br>(-0.572/0.411) | 0.696 |
| ΔKnee angular<br>velocity at RL ($^{\circ}\cdot s^{-1}$) | -0.129 (-0.587/0.392) | 0.634 | -0.149<br>(-0.600/0.374) | 0.582 |
| ΔPeak knee extension<br>velocity ($^{\circ}\cdot s^{-1}$) | 0.089 (-0.425/0.560) | 0.742 | 0.149<br>(-0.374/0.600) | 0.582 |
| ΔKnee angular<br>velocity at TD ($^{\circ}\cdot s^{-1}$) | -0.107 (-0.572/0.410) | 0.693 | -0.082<br>(-0.555/0.431) | 0.762 |

$p^{*}<0.05, p^{**}<0.01$
NDH, Nordic hamstring; MSS, maximal sprint speed; SL, step length; SR, step rate; TD
distance, touchdown distance; RL, distance, release distance; ST, support time; FT, flight
time. RL, Contralateral release. TD, ipsilateral touchdown. Items in bold are those with
significant correlations

Table 4 shows the relationship between ΔMSS and each variable in the NS. There was no significant correlation between ΔMSS and MSS under NS conditions (r=-0.419, p=0.106). There was a significant positive correlation between ΔMSS and the peak combined angle under NS conditions (r=0.560, p<0.05).

**Table 4.** Relationship between the amount of MSS change by condition and each variable at NS conditions.

| Variables in NS | ΔMSS | |
| --- | --- | --- |
|  | Correlation coefficient<br>(Lower/Upper 95 % CI) | *p*-value |
| MSS | -0.419 (-0.757/0.097) | 0.106 |
| SL | -0.323 (-0.706/0.206) | 0.222 |
| SF | -0.050 (-0.533/0.457) | 0.854 |
| TD distance | 0.266 (-0.264/0.673) | 0.319 |
| RL distance | -0.260 (-0.670/0.270) | 0.331 |
| ST | 0.234 (-0.296/0.654) | 0.382 |
| FT | -0.160 (-0.607/0.365) | 0.555 |
| Support distance | -0.128 (-0.587/0.392) | 0.636 |
| Flight distance | -0.349 (-0.720/0.177) | 0.185 |
| Hip angle at RL | -0.099 (-0.567/0.417) | 0.716 |
| Peak hip flexion angle | -0.052 (-0.534/0.456) | 0.849 |
| Hip angle at TD | -0.308 (-0.697/0.222) | 0.246 |
| Knee angle at RL | 0.171 (-0.355/0.614) | 0.528 |
| Peak knee extension angle | 0.271 (-0.259/0.676) | 0.31 |
| Knee angle at TD | 0.017 (-0.483/0.509) | 0.95 |
| Combined angle at RL | 0.207 (−0.322/0.637) | 0.441 |
| Peak combined angle | 0.560 (0.089/0.826) | **0.024\*** |
| Combined angle at TD | 0.329 (−0.199/0.709) | 0.213 |
| Hip angular velocity at RL | 0.268 (−0.263/0.674) | 0.316 |
| Peak hip extension velocity | −0.255 (−0.666/0.276) | 0.341 |
| Hip angular velocity at TD | −0.037 (−0.523/0.468) | 0.893 |
| Knee angular velocity at RL | 0.399 (−0.121/0.747) | 0.126 |
| Peak knee extension velocity | 0.421 (−0.094/0.758) | 0.104 |
| Knee angular velocity at TD | −0.348 (−0.720/0.178) | 0.186 |

*p*\*<0.05
MSS, maximal sprint speed; SL, step length; SR, step rate; TD distance, touchdown distance; RL distance, release distance; ST, support time; FT, flight time. RL, contralateral release. TD, ipsilateral touchdown. Items in bold are those with significant correlations.

## Discussion

The main finding of this study was that static stretching of the hamstrings for 120 s per leg resulted in a significant decrease in MSS via increased touchdown distance and ST, decreased flight distance, decreased knee extension angular velocity at contralateral release, and increased combined angle during the later phase of late swing. Increased hamstring muscle-tendon unit length during the late swing phase and longer touchdown distance in the early stance phase may increase hamstring strain [11, 69–71]. Thus, it is suggested that relatively long-term static stretching of the hamstrings may not only decrease immediately following MSS but may also induce some kinematic changes associated with HSI risk.

Furthermore, the lower the NDH strength, the more significant was the decrease in MSS, increase in ST, decrease in flight distance, and decrease in peak hip extension angular velocity in the SS. Thus, improving NDH strength may help minimize the negative effects of static stretching on the hamstrings.

### Effect of static stretch on hamstrings on MSS and lower limb kinematics

In this study, under SS conditions, the sprint speed at 52.5–60.0 m was lower than in NS conditions. This result differs from that of Kistler et al. [43], who observed a significant speed reduction only at 20–40 m during a 100 m sprint. Kistler et al. [43], also stretched muscles other than the hamstrings, whereas only the hamstrings were stretched in the present study. Thus, this difference in results can be explained by the differences in the stretched parts of the body. The decrease in MSS at SS in this study was caused by an increase in touchdown distance, which was associated with braking impulses in the support phase [72], an increase in ST, which was strongly associated with MSS [1, 2], and a decrease in flight distance. Hip flexion and knee extension were more prominent because the combined angle at the moment of touchdown was greater in SS than in NS, indicating that these kinematic changes were caused by the increased touchdown distance. This may also be because the function of the hamstrings, which decelerates the forward-moving thighs and lower legs during the swing phase and promotes active touch down close to the body [73], was impaired by static stretching. Relatively strong quadriceps contractions can produce angular momentum at the knee joint, which exceeds the mechanical limits of the hamstrings [74].

Therefore, in the present study, in which only the hamstrings were stretched, static stretching may have decreased the hamstring‒quadriceps ratio and promoted knee extension during the late swing phase, which could have negatively affected the mechanics of MSS acquired.

In contrast, Ruan et al. [39] did not observe a decrease in sprint speed or knee flexion and hip extension peak torque after static stretching of the hamstrings in a 10-m sprint. The MSS phase requires more eccentric power from the hamstrings in hip extension and knee flexion than the acceleration phase [45–47]. In addition, compared to the acceleration phase, MSS enhancement requires a higher level of strong ground strike action with early switching of hip flexion-extension and early deceleration of knee extension [2, 45, 75]. Therefore, the MSS phase was more likely to be negatively affected by static stretching of the hamstrings. Based on the above, it may be advisable to refrain from relatively long-term static stretching of the hamstrings only immediately prior to competitions where a high MSS is required.

Static stretching, which reduces the passive stiffness of the hamstrings, also generates knee flexion peak torque at longer hamstring lengths during accelerated sprinting and isokinetic strength measurements [29, 30, 38, 39]. This is a favorable change for HSI prevention [38, 39, 76, 77]. If the SS condition in this study was also adapted to produce peak knee flexion torque with longer muscle length, it can be concluded that there is a partial benefit for HSI risk reduction. However, this study did not calculate lower-limb kinetics, which limited our reference to this aspect of the study. In contrast, knee extension and hip flexion velocity at contralateral release decreased at SS conditions in the present study. Early generation of large knee flexion and hip extension eccentric torques is necessary for early control of faster-moving lower limb motion. Sprinters begin to exert hip extension torque and decelerate their thighs from mid-swing [45]. However, if static stretching suppresses hip extension torque or delays the timing of peak torque generation, early deceleration of the lower limb, which swings forward at a high speed, will not be possible. Therefore, by purposely suppressing the forward swing speed of the lower limb, the demand for fast eccentric contraction and early large-torque exertion at the hamstrings may have been avoided. Thus, static stretching may have the potential to reduce the risk of HSI during the MSS phase by suppressing high-speed hip extension-flexion motion. However, despite the decrease in hip flexion and knee extension velocity in contralateral release, the knee extension velocity was higher at SS conditions than at NS conditions from around 25% late swing, and the combined angle was larger than in NS toward an ipsilateral touchdown. This change is similar to the finding reported by Wolski et al. [78], who showed that repeated sprint fatigue reduced maximal hamstring strength, increased combined angle, and decreased sprint speed, and that increased lengthening stress may be a risk factor for HIS. The increased touchdown distance causes overstride [79, 80] and increase the horizontal braking impulse in the support phase [72]. This early support phase (5% of the support phase) is another phase of increased HSI risk owing to the increased demand for hamstring forces to resist hip flexion and knee extension caused by large ground reaction forces [69, 70]. Thus, it is suggested that static stretching may produce some kinematics during the MSS phase, from the late swing phase to the early support phase, which is unfavorable for HSI risk. In addition, long-term static stretching of the hamstrings only just prior to a sprint, even if performed during the sprint training phase, should be used with caution because it may cause the athlete to learn kinematics that is not desirable for preventing HSI.

### Characteristics of NDH strength and kinematics in sprinters who are prone to acute impairment at MSS due to static stretching

Many previous studies reported no harmful effects of static stretching in highly trained elite athletes [53–58]. In this study, the higher the hamstring strength level, the smaller the decrease in the MSS. NDH reduces HSI risk by extending the fascicle length and increasing knee flexion eccentric torque with a longer hamstring length [23–28]. NDH strength also correlates with lean angle during NDH measurement, with higher NDH muscle strength at larger knee angles [64]. Thus, athletes with low hamstring compliance and eccentric torque in the limb position, where knee extension and hip flexion are promoted, may be unable to adapt to eccentric torque exertion when the combined angle increases in the late swing due to static stretching, resulting in lower MSS. The evidence suggests that participants with a higher ΔMSS had smaller peak combine angles in the NS. This may be a kinematic feature resulting from a failure to accommodate torque exertion at longer hamstring lengths. Therefore, increased NDH strength is recommended as a condition for athletes who require MSS, not only to reduce the risk of HSI during high-speed sprints but also to minimize or eliminate the acute negative effects of long-term static stretching.

### Limitations

The kinematic analysis in this study used a two-dimensional analysis, which may have resulted in less accurate measurements. There were no significant main effects or interactions of the NS and SS measurement order or date on the variables; however, the effects of weather and wind speed could not be completely eliminated. Furthermore, the hamstring compliance of the participants may have influenced the results of this study. However, this study did not assess participants’ hamstring compliance, and the extent to which compliance was altered by static stretching was unclear. In addition, two participants with HSI injuries from the previous year were included in the study. The history of HSI injury may have influenced the results of this study, because the effects on lower limb kinematics and kinetics on the injured leg during maximal sprinting may persist even 5 years after injury [81]. In addition, the mechanics of the late swing phase may be influenced by those of the prior recovery phase [78]. Therefore, further studies are required to broaden the scope of future analyses. In this study, only the MSS phase was analyzed. The findings of this study should be applied because even participants who did not experience a decrease in MSS due to static stretching may have experienced a negative effect on sprint speed during the acceleration phase.

## Conclusion

Under SS conditions, the touchdown distance increased, flight distance decreased, and ST increased as the MSS decreased. In addition, the greater the MSS reduction due to SS, the lower the NDH peak force and torque, and the smaller the peak combining angle under NS conditions. Therefore, athletes with poor hamstring strength and low compliance with long hamstrings should avoid long-term static stretching immediately before high MSS. Under SS conditions, the knee extension velocity at contralateral release was lower, while the combined angle was larger at ipsilateral touchdown. This suggests that long-term static stretching of the hamstring immediately prior to high-speed sprints may induce kinematics that is undesirable for HSI prevention.

## Data Availability

The raw data supporting this article will be made available by the authors without any undue reservation.

## Acknowledgments

None.

